# Latent class analysis identifies distinctive behavioral subtypes in children with fragile X syndrome

**DOI:** 10.1101/2022.05.12.22275013

**Authors:** Walter E. Kaufmann, Melissa Raspa, Carla M. Bann, Julia M. Gable, Holly K. Harris, Dejan B. Budimirovic, Reymundo Lozano, FORWARD Consortium

## Abstract

Fragile X syndrome (FXS) is associated with a characteristic profile of physical and neurobehavioral abnormalities. These phenotypical features are highly variable among affected individuals, which leads to difficulties in developing and evaluating treatments as well as in determining accurate prognosis. The current investigation employed data from FORWARD, a clinic-based natural history study of FXS, to identify subtypes by applying latent class analysis (LCA).

A pediatric cross-sectional sample of 1,072 males and 338 females was subjected to LCA to identify neurobehavioral classes (groups). Input consisted of multiple categorical and continuous cognitive and behavioral variables, including co-occurring behavioral conditions, sleep and sensory problems, measures of autistic behavior (SCQ, SRS-2), and scores on the Aberrant Behavior Checklist revised for FXS (ABC_FX_). Clinically relevant class solutions were further delineated by identifying predictors using stepwise logistic regressions and pairwise comparisons. Following this, classes were characterized in terms of key demographic, genetic, and clinical parameters.

LCA fit parameters supported 2- to 6-class models, which showed good correspondence between patterns of co-occurring conditions and scores on standardized measures. The 5-class solution yielded the most clinically meaningful characterization of groups with unique cognitive and behavioral profiles. The “Mild” class (31%) included patients with attention problems and anxiety but few other major behavioral challenges as reflected by scale scores. Most individuals in the “Severe” class (9%) exhibited multiple co-occurring conditions and high mean scale scores on behavioral measures. Three “Moderate” classes were identified: a “Moderate Behavior” class (32%), a “Social Impairment” class (7%), and a “Disruptive Behavior” class (20%). All classes displayed distinctive SRS-2, SCQ, and ABC_FX_ profiles, which reflected their degree of non-overlap as estimated by pairwise effect sizes. Groups differed with regard to sex, intellectual disability, autism spectrum disorder diagnosis, and medication use.

These findings support the notion that, it is possible to identify behavioral subtypes in children with FXS, reflecting both overall level of severity and specific areas of impairment. These subtypes have implications for clinical management and therapeutic development and assessment. Future studies are needed to determine the stability of these group profiles and their relationship with other aspects of the FXS phenotype.

## Introduction

Fragile X Syndrome (FXS) is the most common inherited neurodevelopmental disorder, affecting up to 1 in 7,000 males and 1 in 11,000 females in the United States (1). FXS is linked to the expansion of a CGG repeat (≥ 200, termed full mutation) in the 5’ untranslated region of the fragile X messenger ribonucleoprotein 1 (*FMR1*) gene. This leads to epigenetic silencing, by atypical DNA methylation, and the consequent reduction in *FMR1*’s protein, FMRP (2, 3). Through its function as a translational regulator, FMRP plays an important role in synaptic development and function. Levels of FMRP correlate with overall severity of the FXS phenotype, with males being more severely affected due to the X-linked pattern of inheritance of *FMR1* mutations (3, 4, 5, 6).

FXS is characterized by a range of physical features, such as large ears, long face and joint laxity, and medical problems of variable frequency (e.g., common recurrent otitis media, rare mitral valve prolapse) (7). However, FXS neurologic and behavioral manifestations are among those with greatest impact on functioning and quality of life (8, 9, 10). Cognitive impairment manifests as intellectual disability (ID) in >90% of males (predominantly moderate ID) and ∼50% of females (mainly borderline-mild ID) 7 years or older, and it is associated with language impairment and other dysfunctions (e.g., deficit in executive function) (11). Common behavioral abnormalities include attention-deficit/hyperactivity disorder (ADHD) symptoms, namely hyperactivity, impulsivity, and attentional difficulties; anxiety; increased sensory reactivity (i.e., hypersensitivity); and autistic behaviors (12, 13). The latter could be severe, leading to the diagnosis of autism spectrum disorder (ASD) in almost 50% of males and 17% of females 3 years or older (11, 14). Other neurologic and behavioral abnormalities include seizures (15), sleep problems (16), perseverative behavior, and disruptive behavior also termed IAAS (irritability/agitation, aggression, and self-injury) (12, 17).

The literature on cognitive and behavioral impairments in FXS supports a distinctive profile, which has helped in the identification of unusual phenotypes as linked to *FMR1* mutations (e.g., Prader-Willi phenotype, mild-moderate ID with prominent social anxiety) (12, 18). However, there is considerable variability in the severity and pattern of association of neurobehavioral features in FXS. A profile characterized by the association of severe ID, IAAS, and severe autistic features has been replicated in multiple studies of each of these clinical manifestations (14, 17). Whether similar combinations of neurologic and behavioral features exist among less-affected individuals is not clear. This is in part due to the high prevalence of ADHD and anxiety symptoms across groups of individuals with FXS, in particular those with milder levels of cognitive impairment. Shortcomings inherent to analyses focused on delineating a few features at a time have also limited the identification of distinctive subphenotypes in FXS. New analytical methodologies have raised hopes of identifying reproducible groups in FXS. s and colleagues (2017) application of topological data analysis, an unsupervised type of multivariate pattern analysis, to longitudinal structural MRI data from young children with FXS led to the identification of two large subgroups with distinctive anatomical, cognitive, adaptive functioning, and autistic behavior severity profiles (19). While the most severe group resembled previously described groups of individuals with marked cognitive impairment and severe disruptive and autistic behavior (14, 17), the findings also demonstrated the potential of using unbiased analyses of large datasets for recognizing combinations of neurobehavioral features.

Intra- and inter-individual variability in neurobehavioral features is not only present in FXS but in most neurodevelopmental disorders. It represents a major challenge for developing and implementing therapies and for measuring their outcomes and, ultimately, for improving quality of life in these disorders (20, 21). This has led to cluster analysis studies aiming at identifying clinically relevant groups in other neurodevelopmental disorders, such as idiopathic ASD and Down syndrome (22, 23, 24, 25, 26). Of relevance to subphenotyping efforts in FXS are the recent studies by Wiggins et al. (2017) and Channell and colleagues (2021), which reported, respectively, four subgroups of young children with ASD and three subtypes of individuals with Down syndrome on the basis of multiple measures of cognition and behavior (25, 27). This more complex multidimensional approach contrasted with earlier work using a single behavioral instrument, such as the Autism Diagnostic Interview-Revised (22), the Aberrant Behavior Checklist-Community (ABC-C) (23), and the Social Responsiveness Scale-2 (SRS-2) (28).

Here, we report on a latent class analysis (LCA) of a large database of children with FXS, from a clinic-based natural history study, which integrated multiple continuous and categorical neurobehavioral variables. This work intended to test the hypothesis that, by applying more advanced clustering methods, it is possible to recognize multiple subtypes of individuals with FXS with distinctive and clinically relevant behavioral profiles. In contrast to previous latent profile analysis (LPA) and LCA studies, we also aimed at generating data for implementation of FXS subtyping into clinical practice and research by delineating FXS groups based on widely used clinical instruments and by determining their level of overlap.

## Methods

### Participants

Participants were part of FORWARD, a multisite, clinic-based longitudinal observational study which collects data through clinician- and parent-reported forms and a few standardized instruments. FORWARD represents the largest resource of clinical and demographic data for the FXS population in the United States (11). Twenty-five FXS specialty clinics across the United States participate in FORWARD. After obtaining Institutional Review Board approval, families are enrolled and data are collected annually, typically at the time of the individual’s scheduled clinical visit.

Analyses in this study were performed on a cross-sectional dataset from a subset of individuals with FXS under 21 years of age from the FORWARD dataset Version 6. Only initial baseline data for each individual was included in the analyses, and age was based on the Clinician Report Form. This resulted in a sample size of 1,410 individuals with FXS,1,072 males and 338 females. FORWARD’s diagnoses of FXS were confirmed by clinicians’ review of a genetic test report indicating an *FMR1* full mutation. The majority of individuals with FXS were White (73%) and non-Hispanic (77%), with those from Black/African American (8%), Asian (4%), or other races (15%) comprising the rest of the sample. Families reported a range of annual incomes, with 25% indicating $50,000 or less, 29% between $50,000 and $100,000, and 35% over $100,000 (8% of families chose not to answer this question). About 36% of families reported an associate’s degree or less, with 31% holding a bachelor’s degree, and 33% with a post-graduate degree.

### Measures

Variables introduced into the statistical models (i.e., input variables) included behavioral diagnoses (i.e., co-occurring conditions) and scores from standardized behavioral instruments and FORWARD ad hoc scales.

#### Co-occurring conditions

The presence (Yes/No) of the following co-occurring behavioral abnormalities was collected from the Clinician Report form: Attention problems; Hyperactivity; Hypersensitivity; Anxiety; Obsessive-compulsive disorder/perseverative behavior; Mood swings/depression; and Irritability/Agitation, Aggression, and Self-injury (IAAS). Additional neurobehavioral variables included presence and severity of sensory problems and sleep problems, both of which were assessed by multiple questions on FORWARD’s Clinician Report form (16). Selected questions were grouped to form Sleep and Sensory problem categories (Supplementary Table 1). Sensory problems and sleep problems scores were computed by taking the means of the applicable items.

**Table 1.** Model fit statistics from the latent profile analysis

|  | LCA Solution – Number of Classes |  |  |  |  |  |
| --- | --- | --- | --- | --- | --- | --- |
|  | 1 | 2 | 3 | 4 | 5 | 6 |
| **Overall Model Fit** |  |  |  |  |  |  |
| AIC | 42014 | 38597 | 37508 | 37121 | 36692 | 36499 |
| BIC | 42156 | 38833 | 37839 | 37546 | 37212 | 37113 |
| Sample-adjusted BIC | 42070 | 38690 | 37639 | 37289 | 36898 | 36741 |
| **Likelihood ratio tests for k vs. k-1 classes (p-values)** |  |  |  |  |  |  |
| Vuong-Lo-Mendell-Rubin test | N/A | < 0.001 | < 0.001 | 0.348 | 0.045 | 0.591 |
| Parametric bootstrapped test | N/A | < 0.001 | < 0.001 | < 0.001 | < 0.001 | < 0.001 |

*AIC* Akaike information criterion, *BIC* Bayesian information criterion

#### Standardized behavioral instruments

Three standardized instruments were administered to assess problem behavior, socio-emotional difficulties, and autistic behavior: the Aberrant Behavior Checklist, Community Edition (ABC-C), revised for FXS (ABC_FX_); the Social Responsiveness Scale, Second Edition (SRS-2), and the Social Communication Questionnaire (SCQ). The SRS-2 and the SCQ are completed once over the course of the 4-year cycle of the FORWARD study, whereas the ABC_FX_ is completed annually. SRS-2 Tscores, raw total SCQ score, and ABC_FX_ subscale scores were calculated as described in previous publications (29, 30).

#### Other clinical variables

Clinicians indicated the presence and level of intellectual disability (ID) according to DSM-5 (2013) criteria: None, Borderline, Mild, Moderate, and Severe/Profound (combined due to small sample sizes) (31). Children showing developmental delays, who were younger than 6 years of age, were placed into a sixth category for (Global) Developmental Delay (31). Other clinical variables included current autism spectrum disorder (ASD) diagnosis, as determined by clinician assessment applying DSM-5 criteria, and current use of psychopharmacological medications or investigational drugs for behaviors (Yes/No) (11, 14, 31). Two types of *FMR1* mosaicism were also recorded: allele size mosaicism (mix of full mutation and premutation alleles) and methylation mosaicism (mix of fully and partially methylated full mutation alleles) (12).

### Statistical analyses

Latent class analyses (finite mixture models) were conducted with a mixture of continuous and categorical indicator variables using Mplus version 8 (32) to identify clusters (classes) of participants. Categorical variables included the presence of the co-occurring behavioral abnormalities mentioned above. Continuous variables included raw scores on standardized measures (SRS-2 T-score, SCQ total score, ABC_FX_ subscale scores [Irritability, Hyperactivity, Socially Unresponsive/Lethargic, Social Avoidance, Stereotypy, Inappropriate Speech]) and FORWARD scales (Sensory problems score, Sleep problems score). Given their variable range, scale scores were standardized using a *Z*-score conversion with a metric of a Mean = 0 (entire sample average) and a standard deviation (SD) = 1 for these analyses.

To identify the best fitting model, 1 to 6 class models were tested. Model fit was first assessed using information criteria, namely Akaike information criterion (AIC), Bayesian information criterion (BIC) and sample size-adjusted BIC, for which lower values indicate better fit. This was followed by likelihood ratio tests (Vuong-Lo-Mendell-Rubin test, parametric bootstrapped test), which compare the fit of the current model to a model with one fewer class. The final model was selected based on a combination of statistical and clinical considerations. After determining the best models and, therefore, the selected number of classes, chi-square tests and analyses of variance (ANOVA) were conducted to compare classes in terms of the following demographic and clinical characteristics: age, sex, ID level, ASD diagnosis, current use of psychopharmacological medications or investigational drugs for behaviors, *FMR1* methylation status, and *FMR1* repeat allele mosaic. In cases where the overall comparison across all classes was significant, we also computed pairwise comparisons (i.e., Fisher’s exact, t-tests) and calculated effect sizes to determine the level of non-overlap between two individual classes for each LCA input variable. We defined adequate FXS subtype separation as a Cohen’s *d* greater than 1.5, which indicates more than 1½ standard deviation difference (33). Further delineation of classes as FXS subtypes included stepwise logistic regression analyses, which were conducted separately for co-occurring conditions and scale scores. These analyses identified predictor variables contributing most to the distinction between two individual classes (Variable Retained in Model Comparing Pairs of Classes). We considered explanatory variables those that met the abovementioned effect size criterion and were also predictors in the stepwise logistic regression analyses. This set of analyses were performed using SAS (Version 9.4, 2013; SAS, Inc., Cary, NC) and effect size online calculators (www.campbellcollaboration.org/escalc/html/EffectSizeCalculator-SMD1.php; www.escal.site/).

## Results

### Latent class analysis (LCA) models

Table 1 displays model fit statistics of the LCA, with the 2to 6-class models having adequate information criterion parameters and likelihood ratio test p-values. The 2- and 3-classs models did not provide significant clinical differentiation and, thus, were not considered further. The 6-class solution did distinguish individuals across the clusters, but three of the six groups had small sample sizes (∼10%). The 4- and 5-class models were examined more closely because of their ability to not only differentiate least and most severe groups of individuals with FXS, but also to identify different groups with intermediate level of severity. The main difference between the 4- and 5-class models was that the latter solution provided three clearly delineated moderate severity behavior groups. Since distinguishing groups in the intermediate range of behavioral problems is important for clinical management and research purposes, we selected the 5-class solution as the main output of our analyses. Data on the 4-class model is included as supplementary material (Supplementary Tables 2,3, and 4).

### Description of LCA classes of children with FXS

Table 2 depicts the distribution of input variables for the entire sample and the 5-class model solution, including the percentage of individuals with FXS presenting with each co-occurring behavioral condition, scores on the standardized instruments, and scores on FORWARD’s Sensory problems and Sleep problems scales for each class. A visual depiction of the 5-class profiles, in terms of frequency of co-occurring conditions and *Z*-score converted scores is shown, respectively, in Figures 1 and 2. The “Mild” class (Class 1, 31% of the sample) was characterized by low to moderate percentages of individuals with co-occurring conditions, with the higher percentages (∼50%) corresponding to Attention problems and Anxiety, two of the core features of FXS. Individuals in the Mild class also had below average Sensory problems and Sleep problems scores, lower autism symptomatology, and low ABC_FX_ subscale scores. In contrast, the “Severe” class (Class 5, 9% of the sample) had the highest percentage of individuals with co-occurring conditions, with the exception of Hyperactivity, and high or highest scores on all standardized behavioral instruments and on the FORWARD scales.

**Table 2.** Distribution of input variables for 5-class solution and entire sample

| Variable | Entire Sample<br>(100%) | Class 1<br>Mild<br>(31%) | Class 2<br>Moderate<br>Behavior<br>(32%) | Class 3<br>Moderate/<br>Social<br>Impairment<br>(7%) | Class 4<br>Moderate/<br>Disruptive<br>Behavior<br>(20%) | Class 5<br>Severe<br>(9%) |
| --- | --- | --- | --- | --- | --- | --- |
| **Categorical Variables (%)** |  |  |  |  |  |  |
| Co-occurring conditions |  |  |  |  |  |  |
| Attention problems | 77 | 51 | 89 | 68 | 94 | 91 |
| Hyperactivity | 60 | 29 | 70 | 37 | 93 | 77 |
| Hypersensitivity | 67 | 29 | 90 | 55 | 87 | 89 |
| Anxiety | 77 | 52 | 87 | 89 | 85 | 94 |
| OCD/perseverative behavior | 51 | 23 | 61 | 45 | 67 | 78 |
| Mood swings/depression | 19 | 6 | 19 | 18 | 22 | 52 |
| IAAS | 50 | 14 | 56 | 42 | 78 | 86 |
| **Continuous variable [mean (SD)]** |  |  |  |  |  |  |
| Sensory problems score | 2.18 (0.69) | 1.56 (0.41) | 2.33 (0.52) | 2.06 (0.56) | 2.66 (0.61) | 2.83 (0.61) |
| Sleep problems score | 1.54 (0.45) | 1.42 (0.40) | 1.48 (0.41) | 1.48 (0.44) | 1.72 (0.46) | 1.82 (0.49) |
| SRS-2 T-score | 72.00 (12.10) | 60.34 (8.58) | 70.08 (9.21) | 80.03 (7.81) | 78.51 (8.20) | 87.11 (9.17) |
| SCQ total score | 14.60 (5.02) | 15.21 (4.48) | 13.78 (4.66) | 17.33 (4.81) | 12.80 (5.06) | 17.01 (5.55) |
| ABC<sub>FX</sub> subscales |  |  |  |  |  |  |
| Irritability | 15.52 (12.72) | 4.23 (4.20) | 11.51 (6.86) | 13.30 (8.20) | 27.52 (8.67) | 36.57 (9.09) |
| Hyperactivity | 10.67 (7.38) | 4.14 (3.89) | 9.13 (4.85) | 8.77 (5.50) | 17.74 (5.29) | 20.24 (4.72) |
| Socially unresponsive/Lethargic | 7.37 (6.41) | 2.67 (2.90) | 4.58 (3.09) | 10.44 (4.74) | 10.45 (3.89) | 21.07 (4.92) |
| Social avoidance | 2.81 (3.16) | 1.10 (1.73) | 1.55 (1.79) | 8.04 (2.31) | 2.60 (2.21) | 7.96 (2.30) |
| Stereotypy | 5.40 (4.76) | 1.86 (2.45) | 4.06 (3.35) | 6.86 (4.10) | 8.34 (4.36) | 12.27 (4.01) |
| Inappropriate speech | 3.89 (3.40) | 1.60 (1.89) | 3.29 (2.64) | 5.19 (3.17) | 5.76 (3.60) | 7.05 (3.64) |
| IAAS Irritability/agitation, aggression, self-injury |  |  |  |  |  |  |

**Figure 1.**
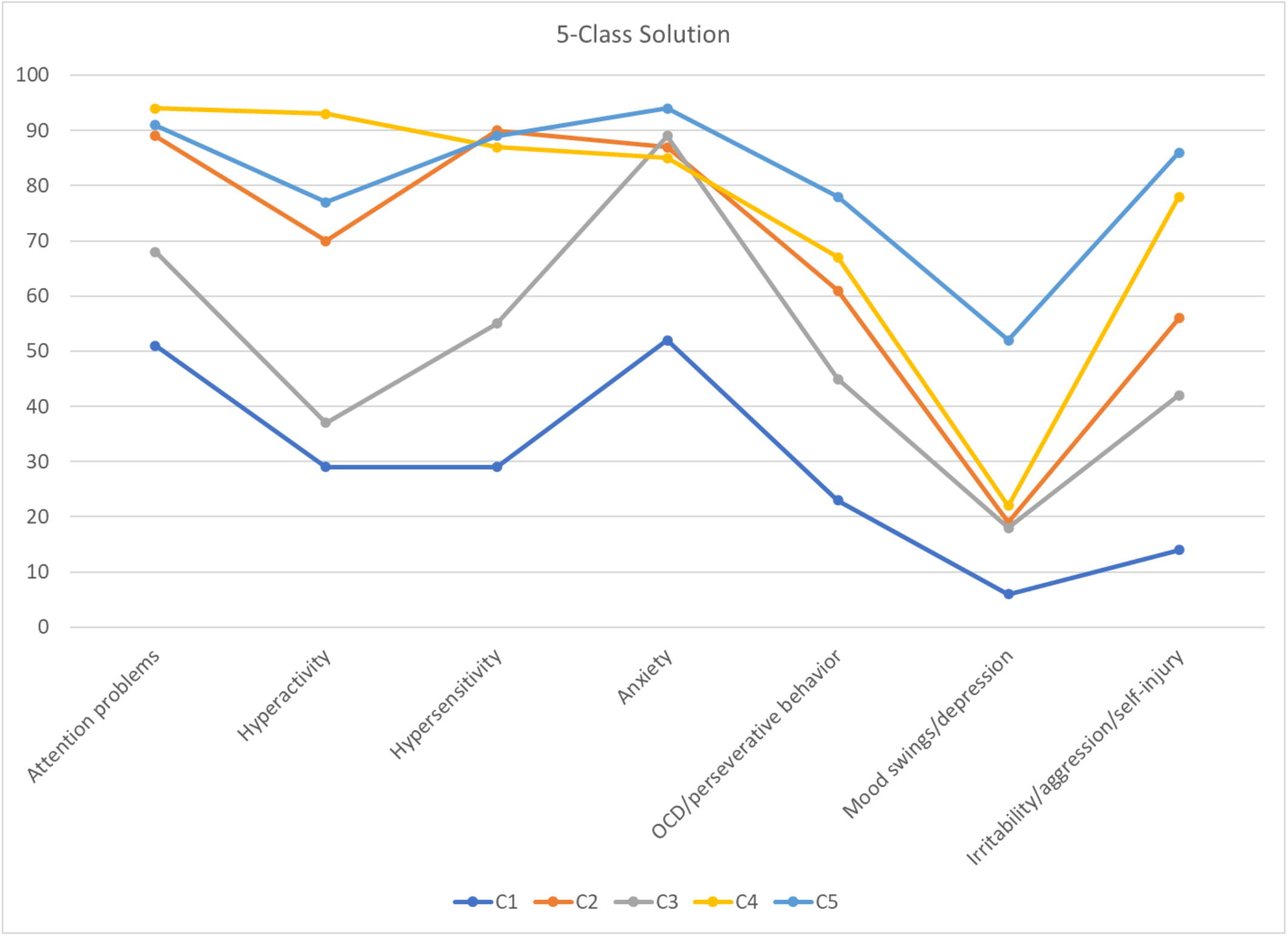
Percentage of respondents by class: co-occurring behavioral conditions in 5-class solution.

**Figure 2.**
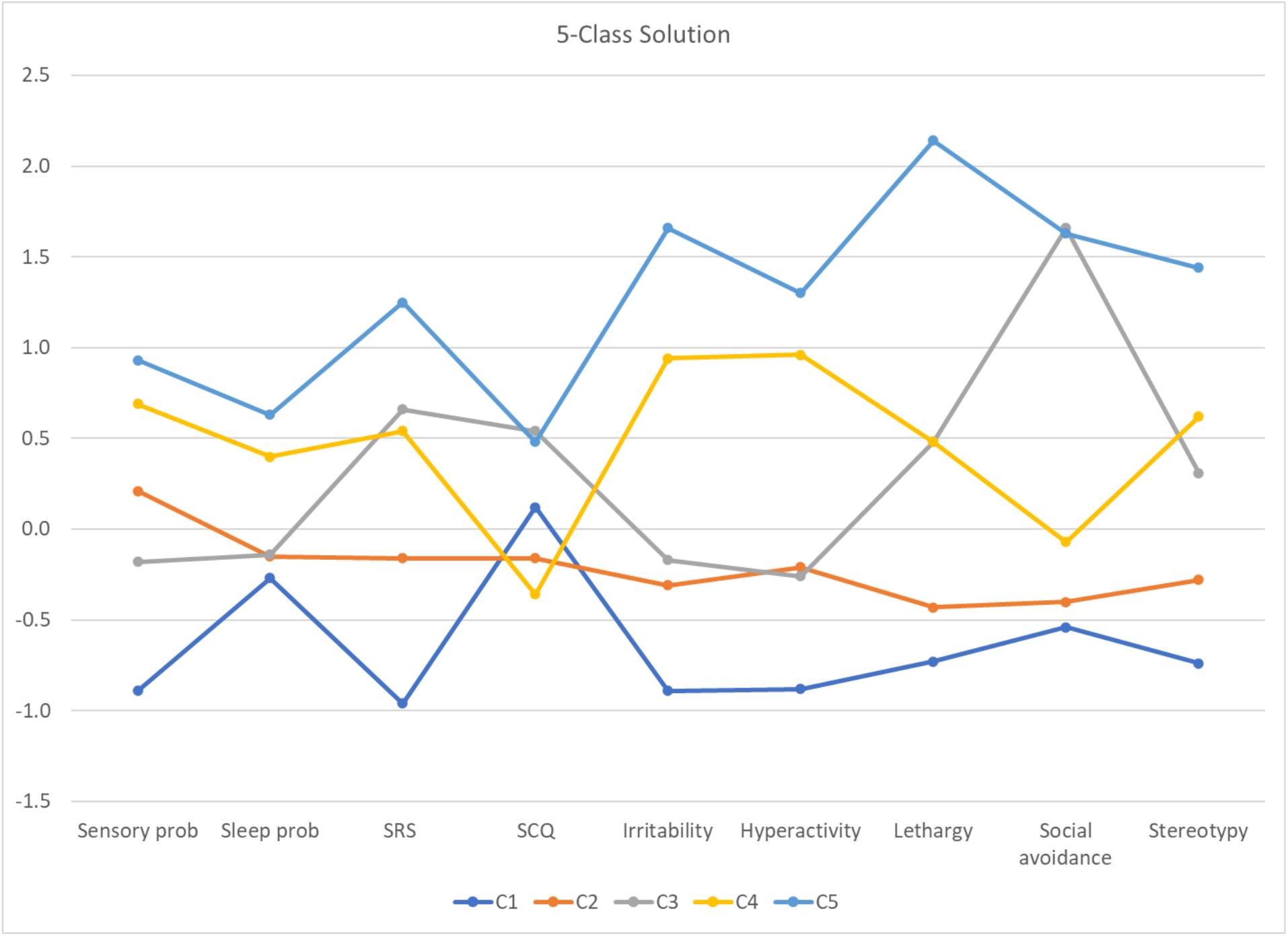
Means by class: standardized scale scores in 5-class solution.

There were three intermediate (Moderate) severity behavior groups, collectively representing approximately 60% of the overall sample. The “Moderate Behavior” class (Class 2, 32% of the sample) had intermediate to high percentages of individuals with ADHD-like symptoms, Hypersensitivity, Anxiety, and IAAS behaviors, and lower scores on the SRS-2, SCQ, and most ABC_FX_ subscales than the other two intermediate groups. The “Moderate/Social Impairment” class (Class 3, 7% of the sample) contained a high percentage of individuals with Anxiety, and those with the highest SRS-2, SCQ and ABC_FX_ Social Avoidance scores among the Moderate groups. The “Moderate/Disruptive Behavior” class (Class 4, 20% of the sample) had the highest percentage of individuals with ADHD-like symptoms, high proportion of IAAS, and high scores on Sensory problems, Sleep problems, and the Irritability, Hyperactivity, and Stereotypy ABC_FX_ subscales.

### Demographic and clinical comparisons between LCA classes

Table 3 depicts clinical characteristics of the FXS subtypes identified in the 5-class solution and the entire sample. As expected, frequency of female subjects and No ID or Borderline ID was the highest in the Mild class and the lowest in the Severe class. Conversely, the predominantly male Severe group had the highest frequency of Moderate and Severe/Profound ID and ASD diagnosis. The Moderate/Social Impairment group was older and had a higher proportion of females than the other two Moderate groups. It also had a slightly higher proportion of individuals with No ID. Frequency of ASD was also significantly different among the Moderate groups, with 55% in the Moderate/Disruptive Behavior group and ∼40% in each of the two other groups. Current use of psychopharmacological medications or investigational drugs for behaviors was distributed according to the range of behavioral impairment of the 5 classes, except for the highest proportion (78%) in the Moderate/Social Impairment group. No statistically significant differences were found between classes in terms of *FMR1* methylation mosaicism; however, the Moderate/Social Impairment group had the lowest proportion with repeat size mosaicism (9% vs. 16-23% in the other groups). Evaluation of level of agreement between frequency of co-occurring conditions and mean scale scores (e.g., proportion with Hyperactivity and ABC_FX_ Hyperactivity scores; proportion with IAAS and ABC_FX_ Irritability scores) was high and assisted by graphical inspection (see Figures 1 and 2).

**Table 3.**
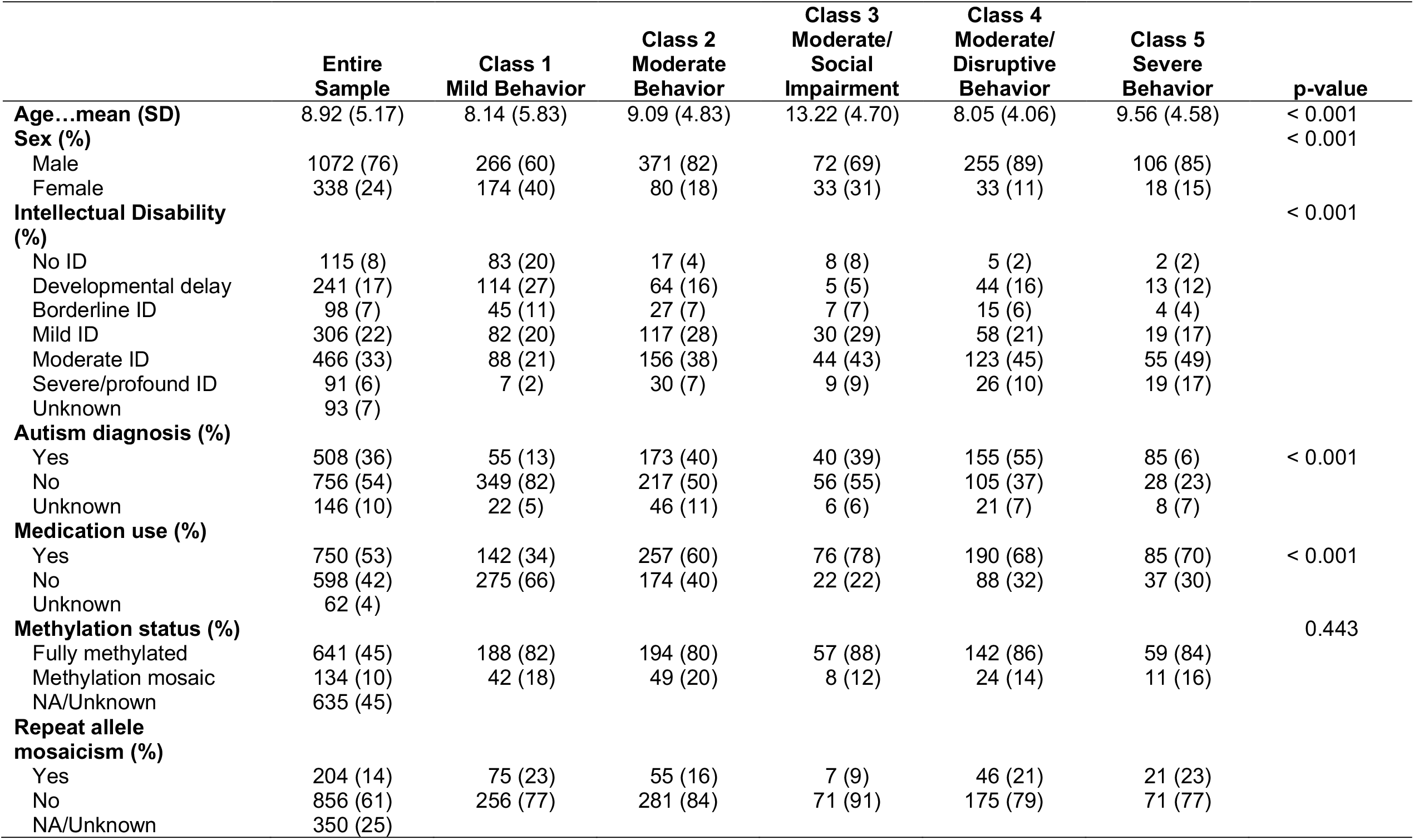
Clinical characteristics of 5-class solution and entire sample

|  | Entire Sample | Class 1<br>Mild Behavior | Class 2<br>Moderate Behavior | Class 3<br>Moderate/<br>Social Impairment | Class 4<br>Moderate/<br>Disruptive Behavior | Class 5<br>Severe Behavior | p-value |
| --- | --- | --- | --- | --- | --- | --- | --- |
| **Age...mean (SD)** | 8.92 (5.17) | 8.14 (5.83) | 9.09 (4.83) | 13.22 (4.70) | 8.05 (4.06) | 9.56 (4.58) | < 0.001 |
| **Sex (%)** |  |  |  |  |  |  | < 0.001 |
| Male | 1072 (76) | 266 (60) | 371 (82) | 72 (69) | 255 (89) | 106 (85) |  |
| Female | 338 (24) | 174 (40) | 80 (18) | 33 (31) | 33 (11) | 18 (15) |  |
| **Intellectual Disability (%)** |  |  |  |  |  |  | < 0.001 |
| No ID | 115 (8) | 83 (20) | 17 (4) | 8 (8) | 5 (2) | 2 (2) |  |
| Developmental delay | 241 (17) | 114 (27) | 64 (16) | 5 (5) | 44 (16) | 13 (12) |  |
| Borderline ID | 98 (7) | 45 (11) | 27 (7) | 7 (7) | 15 (6) | 4 (4) |  |
| Mild ID | 306 (22) | 82 (20) | 117 (28) | 30 (29) | 58 (21) | 19 (17) |  |
| Moderate ID | 466 (33) | 88 (21) | 156 (38) | 44 (43) | 123 (45) | 55 (49) |  |
| Severe/profound ID | 91 (6) | 7 (2) | 30 (7) | 9 (9) | 26 (10) | 19 (17) |  |
| Unknown | 93 (7) |  |  |  |  |  |  |
| **Autism diagnosis (%)** |  |  |  |  |  |  | < 0.001 |
| Yes | 508 (36) | 55 (13) | 173 (40) | 40 (39) | 155 (55) | 85 (6) |  |
| No | 756 (54) | 349 (82) | 217 (50) | 56 (55) | 105 (37) | 28 (23) |  |
| Unknown | 146 (10) | 22 (5) | 46 (11) | 6 (6) | 21 (7) | 8 (7) |  |
| **Medication use (%)** |  |  |  |  |  |  | < 0.001 |
| Yes | 750 (53) | 142 (34) | 257 (60) | 76 (78) | 190 (68) | 85 (70) |  |
| No | 598 (42) | 275 (66) | 174 (40) | 22 (22) | 88 (32) | 37 (30) |  |
| Unknown | 62 (4) |  |  |  |  |  |  |
| **Methylation status (%)** |  |  |  |  |  |  | 0.443 |
| Fully methylated | 641 (45) | 188 (82) | 194 (80) | 57 (88) | 142 (86) | 59 (84) |  |
| Methylation mosaic | 134 (10) | 42 (18) | 49 (20) | 8 (12) | 24 (14) | 11 (16) |  |
| NA/Unknown | 635 (45) |  |  |  |  |  |  |
| **Repeat allele mosaicism (%)** |  |  |  |  |  |  |  |
| Yes | 204 (14) | 75 (23) | 55 (16) | 7 (9) | 46 (21) | 21 (23) |  |
| No | 856 (61) | 256 (77) | 281 (84) | 71 (91) | 175 (79) | 71 (77) |  |
| NA/Unknown | 350 (25) |  |  |  |  |  |  |

### Delineation of LCA classes

Further delineation of FXS subtypes was conducted by comparisons of proportion and mean differences of input variables by chisquare tests (categorical variables) and ANOVA (continuous variables), respectively. Significant overall group differences were followed by pairwise comparisons, with effect sizes included in Table 4. Whereas comparisons between the Mild and Severe classes revealed differences in most input variables, distinction between the extreme groups and the Moderate classes was based on selected variables. Odds ratios from stepwise logistic regression analyses are displayed in Table 5 which provide supportive evidence for a high proportion of variables that were differential between classes in the pairwise comparisons. In general, individual classes were mainly delineated by standardized behavioral measures, which were differential at an effect size (Cohen’s *d*) level of 1.5 or greater. A large proportion of the latter not only met effect size criterion but were also predictors in the stepwise logistic regression analyses. These explanatory variables are bolded in Table 4.

**Table 4.** Effect sizes for pairwise comparisons between classes for 5-class solution (Cohen’s *d*)

|  | 1-2 | 1-3 | 1-4 | 1-5 | 2-3 | 2-4 | 2-5 | 3-4 | 3-5 | 4-5 |
| --- | --- | --- | --- | --- | --- | --- | --- | --- | --- | --- |
| **Categorical Variables** |  |  |  |  |  |  |  |  |  |  |
| Co-occurring conditions |  |  |  |  |  |  |  |  |  |  |
| Attention problems | -1.11<sup>^</sup> | -0.40 | -1.46 | -1.26<sup>^</sup> | 0.71<sup>^</sup> | -0.35 | -0.15 | -1.06 | -0.86 | 0.20 |
| Hyperactivity | -0.94<sup>^</sup> | -0.18 | **-1.87\*<sup>^</sup>** | -1.16<sup>^</sup> | 0.76<sup>^</sup> | -0.93<sup>^</sup> | -0.22 | **-1.69\*<sup>^</sup>** | -0.98<sup>^</sup> | 0.71<sup>^</sup> |
| Hypersensitivity | **-1.69\*<sup>^</sup>** | -0.61<sup>^</sup> | **-1.55\*<sup>^</sup>** | **-1.66\*<sup>^</sup>** | 1.09<sup>^</sup> | 0.14 | 0.03 | -0.94<sup>^</sup> | -1.06<sup>^</sup> | -0.11 |
| Anxiety | -0.98<sup>^</sup> | -1.12<sup>^</sup> | -0.91<sup>^</sup> | -1.42<sup>^</sup> | -0.14 | 0.07 | -0.44 | 0.21 | -0.30 | -0.51 |
| OCD/perseverative behavior | -0.90<sup>^</sup> | -0.55 | -1.04<sup>^</sup> | -1.35<sup>^</sup> | 0.35 | -0.14 | -0.45<sup>^</sup> | -0.49 | -0.80<sup>^</sup> | -0.31 |
| Mood swings/depression | -0.74 | -0.68 | -0.83<sup>^</sup> | **-1.58\*<sup>^</sup>** | 0.06 | -0.09 | -0.84<sup>^</sup> | -0.16 | -0.90<sup>^</sup> | -0.75<sup>^</sup> |
| IAAS | -1.12 | -0.81<sup>^</sup> | **-1.69\*<sup>^</sup>** | **-2.00\*<sup>^</sup>** | 0.32<sup>^</sup> | -0.57<sup>^</sup> | -0.88<sup>^</sup> | -0.89<sup>^</sup> | -1.19<sup>^</sup> | -0.30 |
| **Continuous variables** |  |  |  |  |  |  |  |  |  |  |
| Sensory problems score | **1.64\*<sup>^</sup>** | 1.13 | **2.21\*<sup>^</sup>** | 2.75* | -0.51<sup>^</sup> | 0.59<sup>^</sup> | 0.92 | 1.01<sup>^</sup> | 1.31 | 0.28 |
| Sleep problems score | 0.15 | 0.15 | 0.71 | 0.95 | 0.00 | 0.56<sup>^</sup> | 0.79 | 0.53 | 0.73 | 0.21 |
| SRS-2 T-score | 1.09<sup>^</sup> | **2.35\*<sup>^</sup>** | **2.16\*<sup>^</sup>** | 3.06* | 1.11<sup>^</sup> | 0.95<sup>^</sup> | 1.85* | -0.19 | 0.83<sup>^</sup> | 1.01 |
| SCQ total score | -0.31 | 0.46 | -0.51 | 0.38 | 0.76 | -0.20 | 0.66 | -0.91<sup>^</sup> | -0.06 | 0.81 |
| ABC<sub>FX</sub> subscales |  |  |  |  |  |  |  |  |  |  |
| Irritability | 1.25 | **1.66\*<sup>^</sup>** | **3.52\*<sup>^</sup>** | **5.56\*<sup>^</sup>** | 0.25 | **2.10\*<sup>^</sup>** | 3.39* | **1.67\*<sup>^</sup>** | **2.69\*<sup>^</sup>** | 1.03 |
| Hyperactivity | 1.12<sup>^</sup> | 1.07 | 2.97* | 3.92* | -0.07 | **1.71\*<sup>^</sup>** | 2.30* | **1.68\*<sup>^</sup>** | **2.24\*<sup>^</sup>** | 0.49 |
| Socially unresponsive/Lethargic | 0.64<sup>^</sup> | 2.25* | 2.30* | 5.24* | **1.67\*<sup>^</sup>** | **1.71\*<sup>^</sup>** | **4.64\*<sup>^</sup>** | 0.00 | 2.20* | **2.52\*<sup>^</sup>** |
| Social avoidance | 0.26 | **3.67\*<sup>^</sup>** | 0.76 | 3.63* | **3.40\*<sup>^</sup>** | 0.53 | 3.36* | **-2.43\*<sup>^</sup>** | -0.03 | **2.40\*<sup>^</sup>** |
| Stereotypy | 0.74 | **1.70\*<sup>^</sup>** | 1.88* | 3.57* | 0.80 | 1.13<sup>^</sup> | 2.35* | 0.35 | 1.33 | 0.92<sup>^</sup> |
| Inappropriate speech | 0.72<sup>^</sup> | 1.58* | 1.49 | 2.23* | 0.69 | 0.81<sup>^</sup> | 1.31 | 0.16 | 0.54 | 0.36 |

IAAS Irritability/agitation, aggression, self-injury; \*Cohen's *d* ≥ 1.5; <sup>^</sup>Predictor in stepwise logistic regression models; Bolded, Explanatory variables

**Table 5.**
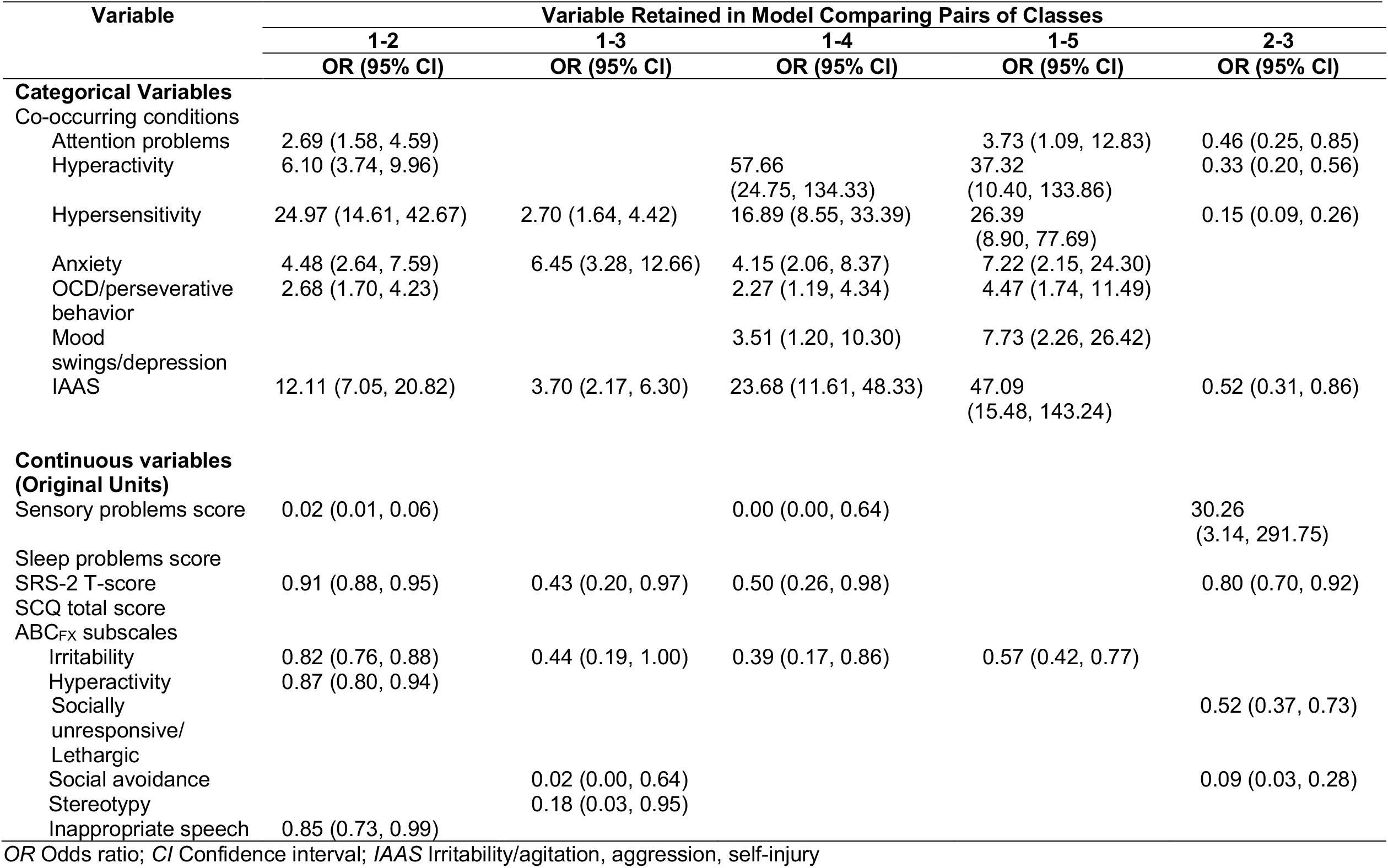
Odds ratios of input variables retained in stepwise logistic regression models of class membership: 5-class solution

Thus, the 5-class solution was characterized by a Mild class with relatively low scores on most standardized measures and FORWARD’s Sensory problems and Sleep problems scales; a Severe class with the opposite pattern, namely highest scores on virtually all scales; a Moderate Behavior class with relatively low scores on all scales with the exception of Sensory problems, in correspondence to its high frequency of Hypersensitivity; a Moderate/Social Impairment class with high scores on the SRS-2, SCQ, and Socially Unresponsive/Lethargic and Social Avoidance ABC_FX_ subscales; and a Moderate/Disruptive Behavior class with high scores on Sensory problems, Sleep problems, SRS-2, and Irritability, Hyperactivity, and Socially Unresponsive/Lethargic ABC_FX_ subscales. These profiles were in correspondence with the frequency of co-occurring behavioral conditions in each class (e.g., low frequency of Hyperactivity, Hypersensitivity, and IAAS in the Mild class; intermediate to high frequency of Hypersensitivity and IAAS in the Moderate Behavior class). Although distinction among the three Moderate classes and between Mild and Moderate Behavior classes were the most difficult, there was at least one set of scale scores that met criteria for explanatory variable and, thus, separated individual classes. Sleep problems and SCQ were the only two subscales that did not differentiate classes using the Cohen’s *d* > 1.5 criterion.

## Discussion

Fragile X syndrome (FXS) is the most common inherited neurodevelopmental disorder. Despite its well-recognized profile of physical and neurobehavioral abnormalities and straightforward diagnosis by genetic testing (12), FXS phenotypical variability represents a challenge for developing and evaluating treatments as well as for determining accurate prognosis (21, 30). Successful subphenotyping in other neurodevelopmental disorders has applied approaches such as LPA and LCA (24, 25, 27). Here, we used LCA (different from LPA in that it also includes categorical variables) for identifying FXS subtypes of clinical relevance in the clinic-based FORWARD natural history database. Cross-sectional analyses incorporated multiple categorical and continuous cognitive and behavioral variables, including scores on standardized instruments (i.e., SCQ, SRS-2, ABC_FX_). Models with 4- and, particularly, 5-class solutions showed the best statistical fit parameters and clinically significance. They identified Mild, Severe and two to three Moderate classes. In the 5class solution, which provided the finest subtype delineation, groups labeled as Moderate Behavior, Moderate/Social Impairment, and Moderate/Disruptive Behavior emerged. Pairwise comparisons of co-occurring behavioral conditions and scale scores, as well as stepwise logistic regression models incorporating these parameters as class predictors, supported the identified FXS subtypes and identified variables for their distinction in clinical and research applications.

The FORWARD project represents a unique resource for FXS clinical research. Despite the limitations of being clinic-based, FORWARD’s database contains a wide range of parameters that can assist in the delineation of meaningful profiles of affected individuals (11, 14, 16, 17, 30). Thus, in contrast with previous subphenotyping studies of neurodevelopmental disorders that used only standardized instruments, we applied LCA using both categorical and continuous variables that included standardized and project-based scales. Mild and severe FXS subphenotypes have been recognized for decades, mainly in the context of *FMR1* mosaicism and female phenotypes and severe autistic behavior and IAAS, respectively (12, 13). More recently, unbiased analytical approaches have solidified these profiles and provided anatomical correlates for more and less severe clinical presentations in FXS (19). However, these broad categories have limited clinical application because of their wide range of behavioral symptom severity.

Major goals of any subtyping effort in FXS include not only identifying mildly and severely affected individuals, but also to delineate groups with intermediate level of behavioral abnormalities. The present study accomplished these objectives by, for the first time, delineating consistent moderate severity groups representing in combination approximately 60% of a FXS pediatric sample. Following their initial identification by LCA, our approach for delineating FXS subtypes combined pairwise class comparisons and logistic regression analyses incorporating LCA input variables. This convergent evidence ensured that the distinctive features of each FXS subtype had solid bases. The adequacy of our strategy was better illustrated by the three Moderate groups in the 5-class solution. Although their cognitive profile was similar, their frequency of co-occurring behavioral conditions and scale scores were distinctive. Class 3, the Moderate/Social Impairment group, had high scores on measures of social behavior but a moderate frequency of ASD (40%) that suggested predominantly social anxiety. This profile is in line with its higher proportion of females, who frequently present with social anxiety (4, 34). Class 4, the Moderate/Disruptive Behavior group, had a profile that resembled that of the Severe group (class 5) but with a lower proportion of ASD diagnosis (55% vs. 85%) and severe/profound ID (10% vs. 17%). These differences among most affected individuals are supported by previously reported profiles of groups of patients with IAAS (17) and ASD (30) in FORWARD. The Moderate Behavior group (Class 2) displayed core behavioral abnormalities of FXS, namely ADHD features, anxiety, and sensory problems in a predominantly male group, which differentiated it from previously reported mildly affected groups with a large proportion of females (4, 12, 34).

Since one of the objectives of the present study was to serve as reference for clinicians and investigators planning to introduce FXS subtyping into their work, we used widely available clinical instruments and statistical parameters for determining level of overlap between groups. For the latter, we employed a conservative effect size of Cohen’s *d* > 1.5 that represents a non-overlap of 71%. Although not all variables differentiating two classes at this effect size level were predictors of subtypes in the stepwise logistic regression analyses, because of their co-variance, a high proportion of them were informative (i.e., explanatory variables) and they should be considered as both distinctive and specific features of FXS subtypes that would allow the identification of individuals in each subphenotype.

The reported FXS subtypes have confirmed and expanded phenotypical profiles associated with the disorder. Among them, the milder features of affected females (4, 12, 34), IAAS as a major component of severe phenotypes (12, 13, 17, 30), and the association between female sex, milder ID, and non-ASD social impairment (12, 13, 34). Our LCA work also supports the core nature of attention problems and anxiety in FXS, since they were present in at least one-third of individuals in every identified class. The application of FXS subtypes to clinical practice and research will ultimately determine their validity and usefulness. However, the present results clearly align with clinical behavioral profiles and, thus, will be useful in the addressing major challenges associated with the disorder. For instance, the clusters can be used to determine the frequency and characteristics of ASD in FXS independent of cognitive impairment. Whether, given the null results examining differences among the clusters by methylation status and allele mosaicism, the identified FXS classes represent distinctive neurobiological entities remains to be demonstrated. However, the topological data analysis reported by Bruno and colleagues (2017) suggests that this may be the case (19).

Despite its important implications, the study also had limitations. They included the use of multiple caregiver-completed behavioral instruments and ad hoc measures for assessing sensory and sleep problems was a shortcoming, which limits the reproducibility of some aspects of the reported profiles. Because of the cross-sectional nature of the study, the potential of these FXS subtypes for informing prognosis will require additional longitudinal analyses. Since our data was exclusively pediatric, stability of FXS subtypes will also need to be determined by incorporating adults into future studies. Finally, the relationship between FXS behavioral subtypes and other aspects of the FXS phenotype, such as physical features, will need to be defined in follow up studies.

## Conclusions

FXS is a phenotypical variable neurodevelopmental disorder, a major limitation for its management and the development of new treatments. Despite the consistent reports of mildly and severely affected groups, comprehensive FXS subphenotyping is lacking. The current pediatric study constitutes the first step in identifying groups across the FXS spectrum of behavioral severity. The proposed FXS subtypes, which can be readily implemented in clinical practice and research, provide the basis for future studies using a precise medicine approach to the disorder.

## Data Availability

De-identified data are available according to the FORWARD project's Data Sharing Plan and the policies of the Centers for Disease Control and Prevention.

## Abbreviations

FXS: Fragile X syndrome
LCA: Latent class analysis
SCQ: Social Communication Questionnaire
SRS-2: Social Responsiveness Scale-2
ABC_FX_: Aberrant Behavior Checklist revised for FXS
FMRP: Fragile X Mental Retardation Protein
(ID): intellectual disability
ADHD: Attention-deficit/hyperactivity disorder
ASD: Autism spectrum disorder
IAAS: Irritability/agitation, aggression, and self-injury
SD: Standard Deviation
AIC: Akaike information criterion
BIC: Bayesian information criterion.

## Supplementary Information

The online version contains supplementary material available at…

## Acknowledgements

We would like to thank everyone involved in the FORWARD project, in particular the participating patients and their families.

## Authors’ contributions

WEK and MR conceived the project and supervised all study efforts. WEK led data analysis and interpretation, and manuscript drafting. CMB had a primary role in data analysis and assisted with data interpretation and drafting the manuscript. JMG helped with data analysis. HKH, DBB, and RL helped with data interpretation and drafting the manuscript. The authors read and approved the final manuscript.

## Availability of data and materials

De-identified data are available according to the FORWARD project’s Data Sharing Plan and the policies of the CDC.

## Declarations

### Ethics approval and consent to participate

The Institutional Review Board at every participating site provided approval and oversight to ensure ethical conduct of human subjects research throughout the study. Informed consent was provided by all parents/guardians who participated. Consent or assent was provided by participants with fragile X syndrome, depending on age and consenting capacity as determined by the Institutional Review Boards.

### Consent for publication

Not applicable.

