## Supplemental Tables for "Latent class analysis identifies distinctive behavioral subtypes in children with fragile X syndrome"

### Supplementary Material

**Supplementary Table 1**
Sleep problem and sensory problem scales

| Item | Item-Total Correlation |
|---|---|
| **Sensory problems*** | |
| Does the child respond too strongly to sensory information in his/her environment? | 0.65 |
| Does the child have signs of hyperarousal? | 0.71 |
| Does the child have unusual sensory input or sensory seeking behaviors? | 0.59 |
| Does the child's sensory problems and hyperarousal restrict participation in everyday activities in the family? | 0.70 |
| **Sleep problems**** | |
| Child does not fall asleep within 20 minutes after going to bed | 0.35 |
| Child struggles at bedtime | 0.47 |
| Child awakens once or more during the night | 0.38 |
| Child seems tired | 0.30 |
| During sleep, child grinds teeth, wets the bed, or appears restless | 0.31 |

*Note: Cronbach's alpha = 0.83
**Note: Cronbach's alpha = 0.60

**Supplementary Table 2**
Distribution of input variables for 4-class solution

| Variable | 4-Class Solution | | | |
|---|---|---|---|---|
| | **Class 1 Mild (32%)** | **Class 2 Moderate Behavior (33%)** | **Class 3 Moderate/ Disruptive Behavior (24%)** | **Class 4 Severe (11%)** |
| **Categorical Variables…%** | | | | |
| Co-occurring conditions | | | | |
| Attention problems | 50 | 90 | 87 | 92 |
| Hyperactivity | 29 | 65 | 83 | 83 |
| Hypersensitivity | 28 | 89 | 80 | 90 |
| Anxiety | 53 | 90 | 82 | 94 |
| OCD/perseverative behavior | 22 | 62 | 60 | 78 |
| Mood swings/depression | 6 | 20 | 19 | 48 |
| IAAS | 15 | 53 | 72 | 89 |
| **Continuous variables…mean (SD)** | | | | |
| Sensory problems score | 1.57 (0.41) | 2.30 (0.52) | 2.55 (0.62) | 2.82 (0.63) |
| Sleep problems score | 1.42 (0.41) | 1.46 (0.39) | 1.69 (0.48) | 1.83 (0.46) |
| SRS-2 T-score | 61.13 (9.25) | 71.03 (9.57) | 78.07 (8.31) | 86.93 (8.73) |
| SCQ total score | 15.29 (4.50) | 14.25 (4.71) | 13.57 (5.33) | 16.38 (5.78) |
| $ABC_{FX}$ subscales | | | | |
| Irritability | 4.33 (4.35) | 10.48 (6.19) | 24.64 (7.90) | 37.61 (8.61) |
| Hyperactivity | 4.17 (3.89) | 8.50 (4.68) | 16.11 (5.44) | 20.74 (4.90) |
| Socially unresponsive/Lethargic | 2.92 (3.19) | 4.85 (3.42) | 10.18 (4.11) | 19.81 (5.45) |
| Social avoidance | 1.35 (2.16) | 2.21 (2.66) | 3.42 (3.09) | 6.93 (3.12) |
| Stereotypy | 1.97 (2.50) | 4.16 (3.34) | 7.73 (4.33) | 12.39 (3.78) |
| Inappropriate speech | 1.64 (1.91) | 3.35 (2.59) | 5.67 (3.54) | 6.94 (3.74) |

*IAAS* Irritability/agitation, aggression, self-injury

**Supplementary Table 3**
Effect sizes for pairwise comparisons for 4-class solution (Cohen's *d*)

| Variable | Effect Sizes for Pairwise Comparisons between Classes | | | | | |
|---|---|---|---|---|---|---|
| | **1-2** | **1-3** | **1-4** | **2-3** | **2-4** | **3-4** |
| **Categorical Variables** | | | | | | |
| Co-occurring conditions | | | | | | |
| Attention problems | -1.19^ | -1.05 | -1.37^ | 0.15 | -0.17 | -0.32 |
| Hyperactivity | -0.85^ | -1.38^ | -1.38^ | -0.53^ | -0.53^ | 0.00 |
| Hypersensitivity | **-1.69*^** | -1.31^ | **-1.75*^** | 0.38^ | -0.06 | -0.44 |
| Anxiety | -1.12^ | -0.76^ | -1.47^ | 0.36^ | -0.35 | -0.71^ |
| OCD/perseverative behavior | -0.95^ | -0.91^ | -1.39^ | 0.04 | -0.44^ | -0.48 |
| Mood swings/depression | -0.73 | -0.71 | -1.45^ | 0.02 | -0.71^ | -0.74^ |
| IAAS | -1.03^ | -1.47^ | **-2.12*^** | -0.45^ | -1.09^ | -0.64^ |
| **Continuous variables** | | | | | | |
| Sensory problems score | **-1.55*^** | -1.92* | -2.62* | -0.44 | -0.95 | -0.43^ |
| Sleep problems score | -0.10 | -0.61 | -0.97 | -0.53^ | -0.91 | -0.30 |
| SRS-2 T-score | -1.05^ | **-1.93*^** | -2.83* | -0.78^ | -1.69* | -1.05^ |
| SCQ total score | 0.23 | 0.35 | -0.22 | 0.14 | -0.43 | -0.51 |
| $ABC_{FX}$ subscales | | | | | | |
| Irritability | -1.13^ | **-3.20*^** | **-5.67*^** | **-2.03*^** | **-3.97*^** | **-1.60*^** |
| Hyperactivity | -1.00^ | **-2.53*^** | -3.95* | **-1.51*^** | -2.59* | -0.88 |
| Socially unresponsive/Lethargic | -0.58 | -1.98* | -4.27* | -1.43^ | -3.74* | **-2.12*^** |
| Social avoidance | -0.35 | -0.78 | -2.26* | -0.42 | -1.70* | -1.13^ |
| Stereotypy | -0.73 | **-1.64*^** | -3.57* | -0.94^ | -2.39* | -1.11^ |
| Inappropriate speech | -0.74^ | -1.42 | -2.07* | -0.76^ | -1.24 | -0.35 |

*IAAS* Irritability/agitation, aggression, self-injury; *Cohen's *d* $\geq$ 1.5; ^Predictor in stepwise logistic regression models; Bolded, Explanatory variables

**Supplementary Table 4**
Odds ratios of Input variables retained in stepwise logistic regression models of class membership: 4-class solution

| Variable | Variable Retained in Model Comparing Pairs of Classes | | | | | |
|---|---|---|---|---|---|---|
| | 1-2 | 1-3 | 1-4 | 2-3 | 2-4 | 3-4 |
| | OR (95% CI) | OR (95% CI) | OR (95% CI) | OR (95% CI) | OR (95% CI) | OR (95% CI) |
| **Categorical Variables** | | | | | | |
| Co-occurring conditions | | | | | | |
| Attention problems | 4.01 (2.37, 6.81) | | 3.26 (1.04, 10.18) | | | |
| Hyperactivity | 4.59 (2.86, 7.38) | 12.67 (7.56, 21.24) | 43.67 (12.48, 152.89) | 2.51 (1.74, 3.61) | 2.77 (1.65, 4.65) | |
| Hypersensitivity | 21.17 (12.83, 34.92) | 8.55 (5.20, 14.07) | 24.30 (8.68, 68.03) | 0.46 (0.29, 0.71) | | |
| Anxiety | 6.26 (3.65, 10.74) | 3.46 (2.03, 5.89) | 6.69 (2.15, 20.82) | 0.57 (0.37, 0.89) | | 3.29 (1.50, 7.21) |
| OCD/perseverative behavior | 2.87 (1.84, 4.49) | 2.32 (1.41, 3.82) | 3.94 (1.56, 9.92) | | 1.76 (1.09, 2.82) | |
| Mood swings/depression | | | 6.06 (1.68, 21.88) | | 2.97 (1.91, 4.63) | 2.89 (1.86, 4.49) |
| IAAS | 7.91 (4.73, 13.21) | 13.80 (8.23, 23.16) | 48.03 (16.34, 141.18) | 2.32 (1.68, 3.22) | 5.81 (3.30, 10.26) | 2.48 (1.37, 4.49) |
| **Continuous variables (Original Units)** | | | | | | |
| Sensory problems score | 0.03 (0.01, 0.07) | | | | | 0.05 (0.00, 0.65) |
| Sleep problems score | | | | 0.00 (0.00, 0.02) | | |
| SRS-2 T-score | 0.91 (0.88, 0.94) | 0.64 (0.44, 0.92) | | 0.66 (0.50, 0.87) | | 0.59 (0.38, 0.91) |
| SCQ total score | | | | | | |
| $ABC_{FX}$ subscales | | | | | | |
| Irritability | 0.84 (0.79, 0.90) | 0.61 (0.42, 0.87) | 0.59 (0.45, 0.76) | 0.30 (0.15, 0.59) | 0.66 (0.58, 0.75) | 0.41 (0.23, 0.73) |
| Hyperactivity | 0.86 (0.79, 0.93) | 0.45 (0.22, 0.92) | | 0.20 (0.08, 0.51) | | |
| Socially unresponsive/Lethargic | | | | 0.17 (0.07, 0.46) | | 0.27 (0.12, 0.64) |
| Social avoidance | | | | | | 0.32 (0.14, 0.73) |
| Stereotypy | | 0.49 (0.28, 0.88) | | 0.24 (0.11, 0.56) | | 0.39 (0.22, 0.69) |
| Inappropriate speech | 0.83 (0.72, 0.95) | | | 0.47 (0.30, 0.71) | | |

*OR* Odds ratio; *CI* Confidence interval; *IAAS* Irritability/agitation, aggression, self-injury
